# A discrete-time-evolution model to forecast progress of Covid-19 outbreak

**DOI:** 10.1101/2020.06.12.20129981

**Authors:** Evaldo M. F. Curado, Marco R. Curado

## Abstract

Based on well-known infection models, we constructed a new model to forecast the propagation of the Covid-19 pandemic which yields a discrete-time evolution with one day interval. The proposed model can be easily implemented with daily updated data sets of the pandemic publicly available by many sources. It has only two adjustable parameters and is able to predict the evolution of the total number of infected people in a country for the next 14 days, if parameters do not change during this time. The model incorporates the main aspects of the disease such as the the fact that there are asymptomatic and symptomatic phases (both capable of propagating the virus), and that these phases take almost two weeks before the infected person status evolves to the next (asymptomatic becomes symptomatic or symptomatic becomes either recovered or dead). One advantage of the model is that it gives directly the number of total infected people in each day (in thousands, tens of thousands or hundred of thousands). The model was tested with data from Brazil, UK and South Korea, it predicts quite well the evolution of the disease and therefore may be a useful tool to estimate the propagation of the disease.

## 1 Introduction

A newly identified coronavirus capable of infecting humans was found in patients hospitalized in Wuhan, China [1] (and, possibly, in Europe [2]) in December 2019. Named Severe Acute Respiratory Syndrome Coronavirus-2, or SARS-CoV-2, and causing the respiratory syndrome known as Covid-19, it joins a group coronaviruses which originated from animals and resulted in human diseases, including Severe Acute Respiratory Syndrome (SARS) and Middle East Respiratory Syndrome (MERS). However, while these diseases share to some extent similar clinical symptoms, including fever, dry cough and dyspnoea [3], the Covid-19 outbreak quickly spread worldwide and became a global public health crisis. In 30 January 2020 WHO declared Covid-19 disease a Public Health Emergency of International Concern (PHEIC) [4], and soon thereafter, on 11 March 2020 it was declared a global pandemic. Notably, while on 30 January 2020 there were 8.324 confirmed cases worldwide, on 28 May 2020 this number increased to almost 6 million [5]. Economically, it is expected that the Covid-19 pandemic results a recession at least as severe as the 2009 crisis [6]. As such, to reduce the social and economic burden caused by the pandemic, the development of reliable models to forecast its propagation is a promising strategy to assist public policies aiming at controlling further dissemination of the disease.

Many models of propagation of diseases are available, including SIR [7], SEIR, SIS and others [8, 9, 10]. These are powerful models, as they are relatively simple and perform well on many infectious diseases. However, these models are based on differential equations and their implementation with available data are rather challenging, what makes them difficult to be widely understood and implemented. Notably, for Covid-19 the accessible data set is extensive and updated in a daily basis, including number of confirmed cases, deaths, and recovered people per country. This increases the relevance of simple models that can be easily implemented and fed with most current data from the pandemic, facilitating tracking of its propagation and assessment of the efficacy of public policies aiming to control it.

The data set of the pandemic can be found in many sources, including governmental, e.g., the European Centre for Disease Prevention and Control (ECDC) [11], and non-governmental, private sources, e.g. the Johns Hopkins University [5] (JHU, from USA), and the website Worldometer [12]. These sources commonly present small differences in their data sets, but as common characteristics all present daily updated data and, among other quantities, the total number of confirmed cases of infected people and the total number of deaths per country. Here, we present a model that can easily incorporate the available data sets and forecast the number of confirmed cases by Covid-19 in any given country for the next 14 days.

## 2 A discrete-time model for the Covid-19 outbreak

The main points about the pandemic Covid-19 taking into account in our model are the following:

- In a population, we can distinguish five types (classes) of people. They are:
  a. people immune to the virus.
  b. people susceptible (*S*) to the virus but not infected.
  c. people infected but still asymptomatic (*A*). They can contaminate other people but present no symptoms. This phase takes roughly two weeks before clinical symptoms become apparent.
  d. people already presenting the clinical symptoms of the infection (*I*). This phase takes roughly two weeks before either recovery or death.
  e. people recovered (*R*) from the disease or dead.
- The disease follows the sequence: *S*→ *A*→ *I*→ *R*. The time interval, *T*, going from *A*→ *I* commonly vary around two weeks and we will take, for simplicity, *T* = 14 days. The time interval going from *I → R* can also vary from 10 to more than 20 days and again, for simplicity, we will take the same value *T* = 14 days. Clearly, this is a crude simplification since these time intervals can significantly vary per individual. For example, in patients where lungs are severely affected the recovery period is expected to take considerably longer than for patients presenting symptoms of a mild flu only. Therefore, putting an average value *T* = 14 for both phases *A* and *I* is a simplification that can only be supported by fitting with the real data set: if the fitting produces a reasonably accurate forecast, then this average value can be kept. Otherwise, this average value for *T* may be fine tuned by more sophisticated generalizations of the model. Nevertheless, in all cases we have studied, the average error rate of tested predicted values were lower than 5%.
- Immune people are not included in the model as they are assumed to not be affected by the virus, nor transmit it. Furthermore, it is assumed that a recovered person does not become susceptible again, i.e., after phase *I* the person is assumed to not be vulnerable to be re-infected by SARS-CoV-2.

We propose that the daily evolution of Covid-19 can be modelled by the following discrete-time equations:

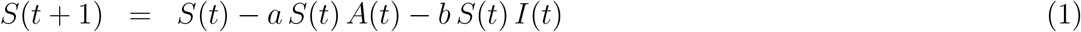

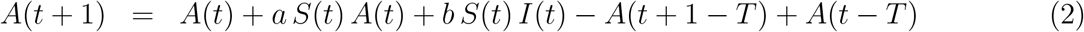

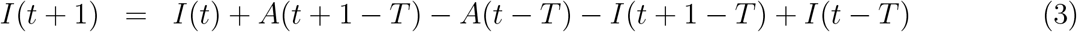

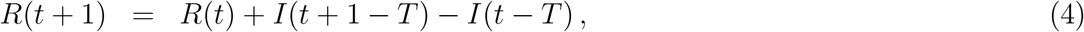

where *t* means the day, *T* means the duration of asymptomatic and symptomatic infected phases, both herein assumed to be 14 days. The term *a S*(*t*) *A*(*t*) indicates that a number of the susceptible people becomes asymptomatic if in contact with asymptomatic people; the same with the term *b S*(*t*) *I*(*t*) where susceptible people becomes asymptomatic if in contact with symptomatic infected people, from now on simply called infected people. The parameters *a* and *b* are constants, measuring the intensity of contagion coming from asymptomatic or infected people and having, in principle, different values. Finally, it is important to note that the sum *S*(*t*) + *A*(*t*) + *I*(*t*) + *R*(*t*) is a constant value and does not depend on time. In fact, it accounts for a fixed part of the studied population, of the order of tens of millions or eventually more for countries with large populations. As such, for these countries, the model may be described in a simplified manner if we consider that *S* is in practical terms a constant (see next section).

The parameter *T* is a key component of the model as it indicates the influence of one class into another. For example, the difference of the number of infected people from time *t* to time *t* + 1, *I*(*t* + 1) −*I*(*t*), see equation (3), is consequence of two factors. The first one is the change in the number of asymptomatic people from time *t* − *T* to *t* + 1 −*T, A*(*t* + 1 −*T*) −*A*(*t* −*T*), because after a time *T* these asymptomatic people turns out to be infected people. The second one is that we have to discount the number of people leaving the class of infected ones from *t* − *T* to *t* + 1 −*T*, because they have more than 14 days since the symptoms of the disease manifest, i.e., *I*(*t* + 1 −*T*) −*I*(*t* −*T*). These number of people are going to the class of recovered (or dead) people, *R*.

## 3 A simplified discrete time model

The model presented in the last section can still be simplified for countries with large population. First, as remarked before, it should be noted that the number of susceptible people can be a large number for countries with millions of inhabitants. The epidemic starts with a very small number of infected people, that increases and can reach (as currently seen) hundreds of thousands of people. For countries with large population, e.g., several tens (or even more than a hundred) million habitants, we can further simplify the model described in equations (1-4) assuming that the number of susceptible people is practically constant in time, *S*(*t*) ≃*S*, where *S* is a constant, so there is no time evolution for *S*. This can be assumed for countries with large populations as the difference between infected (asymptomatic *A* + symptomatic *I*, which theoretically may reach up to few hundred thousand people in a given country) and susceptible people *S* (depending on the country reaching over a hundred million people) is still so huge that the decrease in the sample of susceptible people *S* may be negligible. As such, calling 1 + *a S* ≡ *α* and *b S* ≡ *β*, where *α* and *β* are positive constants, the equations for Covid-19 for large-population countries can be written as:

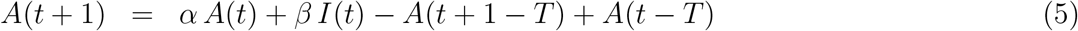

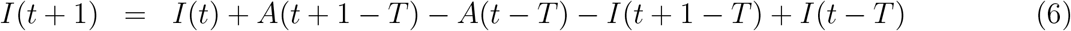

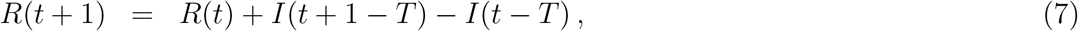

where *A, I* and *R* are counted by hundreds, thousands or hundred of thousands. Hence, here the sum *A* + *I* + *R* is not a constant anymore, but an increasing function on time. The susceptible people are, in these cases, considered as a ‘thermostat’, practically not changing in time. Notice that here the total number of inhabitants of a country is not an important parameter, as it is in other disease models.

This simplified model can be used while the number of *A*+*I* +*R* is lower than a small percentage of the total number of susceptible people of that country. Conservatively, we recommend to use it while this sum is lower than 5%. However, this threshold of 5% is not strict and may be modified to some extent, but if the number of *A* + *I* + *R* is larger than 10% of susceptible people, the accuracy of the simplified version of the model may be affected. In these cases, the complete model represents a better alternative to forecast progress of Covid-19.

An analysis of these equations shows that equations (5-6) are the key equations of the model, with the equation (7) being a supportive equation. This system of equations has only two parameters, *α* and *β*, that can be found phenomenologically for each country and roughly measures the capacity of contagion of asymptomatic and symptomatic people. This model could have more parameters, but it seems that all other parameters can be condensed in these two, with the only requirement of having positive values.

## 4 How to use available data on this model

The data set provided by the above-mentioned sources are updated in a daily basis and have at least two useful information for the proposed model: the number of deaths and the total number of confirmed cases (total symptomatic infected) people,*I*_*tot*_(*t*) per country and globally. The main variables in our model are the number of infected (i.e., asymptomatic *A*(*t*) + symptomatic *I*(*t*)) people at time *t*, i.e. people that at time *t* can infect susceptible people. There are many definitions of symptomatic people in these sources, and they are not completely equivalent. For our model, we will define the class of symptomatic people by means of the total number of confirmed cases by country, as the definition of confirmed cases is the same on all sources.

With our hypothesis we can simply define:

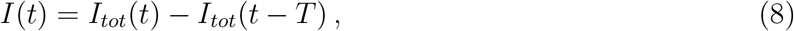

that is, the number of infected people at time *t*, defined as *I*(*t*), is the total number of infected people at time *t* minus the total number of infected people at time *t* − *T*. In turn, the total number of infected people at time *t* − *T* represents people who either recovered or died after the infection, since their symptoms manifested in more than *T* days. So, from the available data set of total infected people from time *t* = *t*_0_, the first day when someone presented the symptoms of the infection, (*I*(*t*_0_) is the number of the first infected people in a particular country at day *t*_0_), until time *t* = *t*_*N*_, the last day where data were collected, we can construct the corresponding data set of the variable *I*(*t*) from *t* = *t*_0_ until *t* = *t*_*N*_. We are assuming, of course, that *I*_*tot*_(*t*_0_− *τ*) were *τ* is a positive integer is equal to zero since we are considering *t*_0_ as the day of the first infected (with symptoms) people. This ensues that *I*(*t*) = 0 for 1≤ *t < t*_0_. In fact, in our model, we always have *t*_0_ = *T* + 1 = 15. Our day one for a specific country is always *t*_0_ −*T*, the day of the first asymptomatic people.

Now, we can construct, from the recent built *I*(*t*) time series, the time series of the other variable, *A*(*t*). For that we use equation (6) rewritten as,

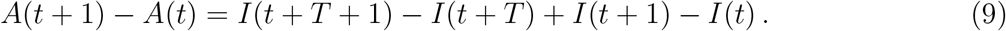

With this equation we can construct the time series of *A*(*t*) from *t* = *t*_0_*>*− *T* = 1 until *t* = −*t*_*N*_ *T*. As *t*_0_ −*T* = 1, i.e., the first day when people got contaminated (but still asymptomatic), we can define the initial condition in order to solve equation (9),

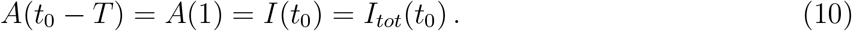

After *T* days these people will lead to the first infected people, at time *t* = *t*_0_ = *T* + 1. So, from the real data set of total infected people we can construct the time series of *I*(*t*) from *t* = 1, *…, t*_*N*_ and, from equations (9) and (10), the time series of *A*(*t*) from *t* = 1, *…, t*_*N*_ −*T*.

In the next step we use equations (5) and (6) to generate the *theoretical* time series for *A*(*t*), from *t* = *t*_*N*_ − *T* + 1, and for *I*(*t*) from *t*_*N*_ + 1. For that we need the values of *α* and *β*.

### 4.1 Obtaining adequate values for *α* and *β*

We need the entire series of *A*(*t*) and *I*(*t*) to forecast the next *T* days for the total number of infected people starting on day *t*_*N*_. One way to try to get good values for the parameters *α* and *β* is to make the prediction using equations (5) and (6) not from *t*_*N*_ but, say, from *t*_*N*_ − 5 until *t*_*N*_, that is, we consider the real data until the day *t*_*N*_ − 5 for *I*(*t*) and the real data until *t*_*N*_ − *T* − 5 for *A*(*t*). This way it is possible to check the theoretical values of *A*(*t*), from *t*_*N*_ −*T*− 5 until *t*_*N*_ −*T, I*(*t*) and *I*_*tot*_(*t*) from *t*_*N*_ − 5 until *t*_*N*_ with the real ones, obtained from the real data set until *t*_*N*_. Adjusting *α* and *β* to best fit these, say, five values, the forecast of the values of *A*(*t*), *I*(*t*) and *I*_*tot*_(*t*) from *t*_*N*_ until *t*_*N*_ + *T* + 1 ensue. In fact, the prediction for *A*(*t*) and *I*(*t*) has no upper bound for the chosen values of *α* and *β*, only the prediction for *I*_*tot*_(*t*) is limited to the next *T* days due to equation (8) if we want to keep using real data. Other ways to obtain good values for the parameters *α* and *β* can be conceived, but we propose this simple way as it worked well in the tested cases.

The values of *α* and *β* can change over time following changes on social behaviour, e.g. due to measures of confinement by local governments or increasing consciousness of the population, which in turn affect the interactions of susceptible people with asymptomatic and symptomatic infected people. Therefore, it is recommendable to test the adequacy of the parameters *α* and *β* along the progression of the disease. In the next section we will see examples of the application of the model in some countries.

## 5 Case studies

If we regard the graphs of total infected people for several countries, we roughly can notice three standard patterns. The first one shows a curve increasing faster than a linear one. These are the cases of Brazil, Russia, India, among others. The second pattern shows an almost linear increasing curve, e.g., UK and US. Finally, the third one presents a strong increasing in the beginning and then the curve tends to saturate, e.g., South Korea, Germany and China. We will discuss examples of each one of these three patterns. In all cases we are considering the data until May 28th. Then, we use the days from May 23th or 24th until May 28th to adjust good values for *α* and *β*. These values can then be used for prediction from May 28th until June 11th.

### 5.1 Brazil

The first infected person in Brazil was identified on February 26th, that is defined here as day *t*_0_. Using *T* = 14, the first contaminated person, according with our model, was identified on February 12th (i.e., 14 days before) defined here as day *t* = 1. The time series of the total infected people from the day 01, February 12th, until May 28th (107 days in total), can be obtained from the sources [5, 11, 12]. We are using herein, in this case, the data set from [5]. Based on this data set we can construct the time series of *I*_*BR*_(*t*) from *t* = 1, *…, t*_*N*_ = 107 and *A*_*BR*_(*t*) from *t* = 1, *…, t*_*N*_ −14 = 93, see Figure 1. Here, we can implement our proposed model to predict 14 days after day 107.

**Figure 1:**
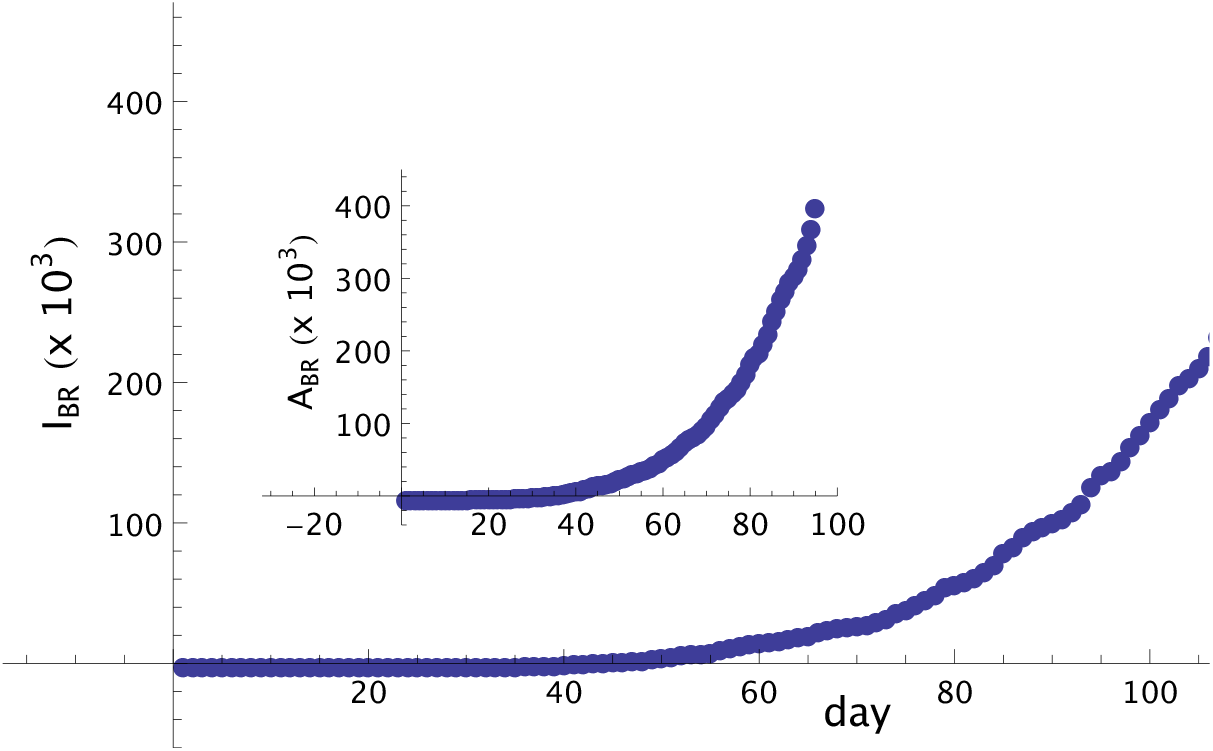
Number of symptomatic infected people in Brazil from day 01, February 12th, until day 107, May 28th. Notice that the curve for symptomatic infected people presents an increasing monotonic behavior. In the inset we show the curve of asymptomatic people until day 93, that also presents a strong monotonically increasing behavior.

With the intention to test the method and find good values for the parameters *α* and *β*, we considered our fictitious 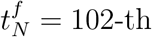 day, so it is possible to compare the total number of infected people as provided by the original source (JHU) with the predicted data generated by the model. For this comparison we will focus on days 102 to 107 and we are considering only the real data of *I*_*BR*_(*t*) up to the day 102 and data of *A*_*BR*_(*t*) up to day 88. Adjusting the predicted data from the method with the real data values, we found that the agreement among the real data and the forecast given by the method from day 102 until day 107 is quite good for *α* = 1.01 and *β* = 0.17, showing that the method, in spite of its simplicity, has good predictive power. The relative error in this period of five days is less than 2%.

This procedure can then be applied to estimate the total number of infected people *T* = 14 days following day 107, see Table 1 and Figure 2. It should be noted that the case of Brazil is special, since these values for the parameters *α* and *β* are practically constant since 30 days before.

**Table 1:** Forecast of the total number of infected people by Covid-19 in Brazil from day 108 (May 29th) until day 121 (June 11th), see also Figure 2. Numbers are in thousands. This forecast was based on values of *I*_*BR*_ and *A*_*BR*_ until day 107 obtained from real data.

|  |  |  |  |  |  |  |  |  |  |  |  |  |  |  |
| --- | --- | --- | --- | --- | --- | --- | --- | --- | --- | --- | --- | --- | --- | --- |
| day | 108 | 109 | 110 | 111 | 112 | 113 | 114 | 115 | 116 | 117 | 118 | 119 | 120 | 121 |
| $I_{tot}(t)$ | 453 | 473 | 499 | 521 | 543 | 568 | 595 | 624 | 658 | 690 | 729 | 767 | 807 | 845 |

**Figure 2:**
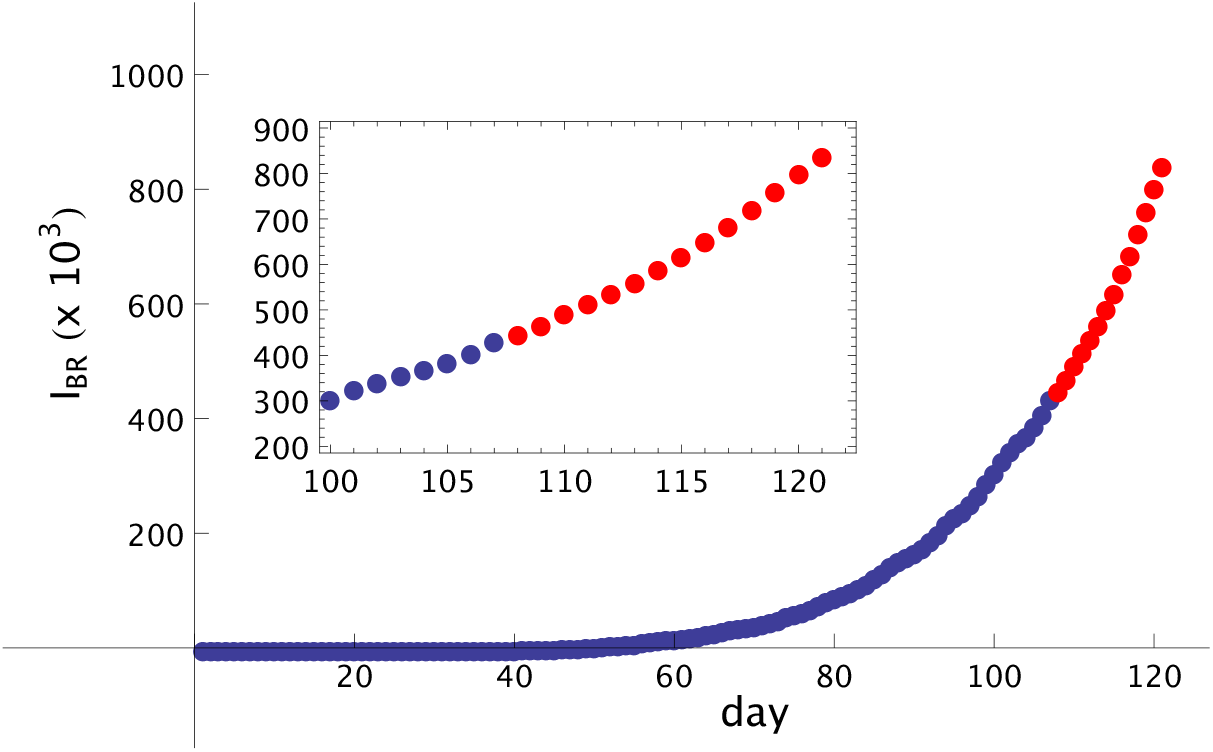
Total number of Covid-19 infected in Brazil as function of the number of days, starting from February 12th, indicated as number 1. Full circles (blue) are real data and empty triangles (red) are the forecast from day 107, May 28th, until day 121, June 11th. *α* = 1.01 and *β* = 0.2. Notice that the curve increases faster than a linear one. In the inset we show a shorter interval, from day 100 up to day 123.

This does not happen with most other countries, where the values of the parameters decrease with time. Accordingly, parameters *α* and *β* should be updated for each change on the monotony of the curve of infected people, since these changes may reflect new social behaviours affecting the contagion parameters *α* and *β*, with consequences on the time evolution of the disease.

### 5.2 UK

Here, we consider the real data of the total number of infected people by Covid-19 in UK. The first people identified in UK presenting symptoms of Covid-19 were registered on January 31th, which we define here as day *t*_0_. Using *T* = 14, the date of contagion for those people was on January 17th, defined here as day *t* = 1. The data set of the total infected people from the day 01, January 17th, until May 28th (133 days in total), can be obtained from the sites [5, 11, 12]. Here we are using the data set from [5]. From this data set we can construct the time series of symptomatic infected people 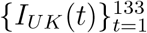, and asymptomatic infected people 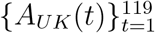, see Figure 3. We note that, in spite of the fact that the graph of total infected people in UK is monotonically increasing, see Figure 4, the graph for symptomatic infected people and asymptomatic people, Figure 3, have a maximum and then decrease, a behavior unlike the behavior of total infected people and clearly pointing out that the peak of the disease has passed by. Besides that, there are many oscillations, increasing and decreasing in a short time. The prediction in a situation like that, with changement of behavior, is more difficult than in the Brazil’s case, where the behavior of total and symptomatic infected people are monotonically increasing. In order to catch this changement of slope, and estimate good values of the parameters *α* and *β* with the intention of forecast, it is better to consider a small interval of time for the test, with all points belonging to the slope of the new tendency.

**Figure 3:**
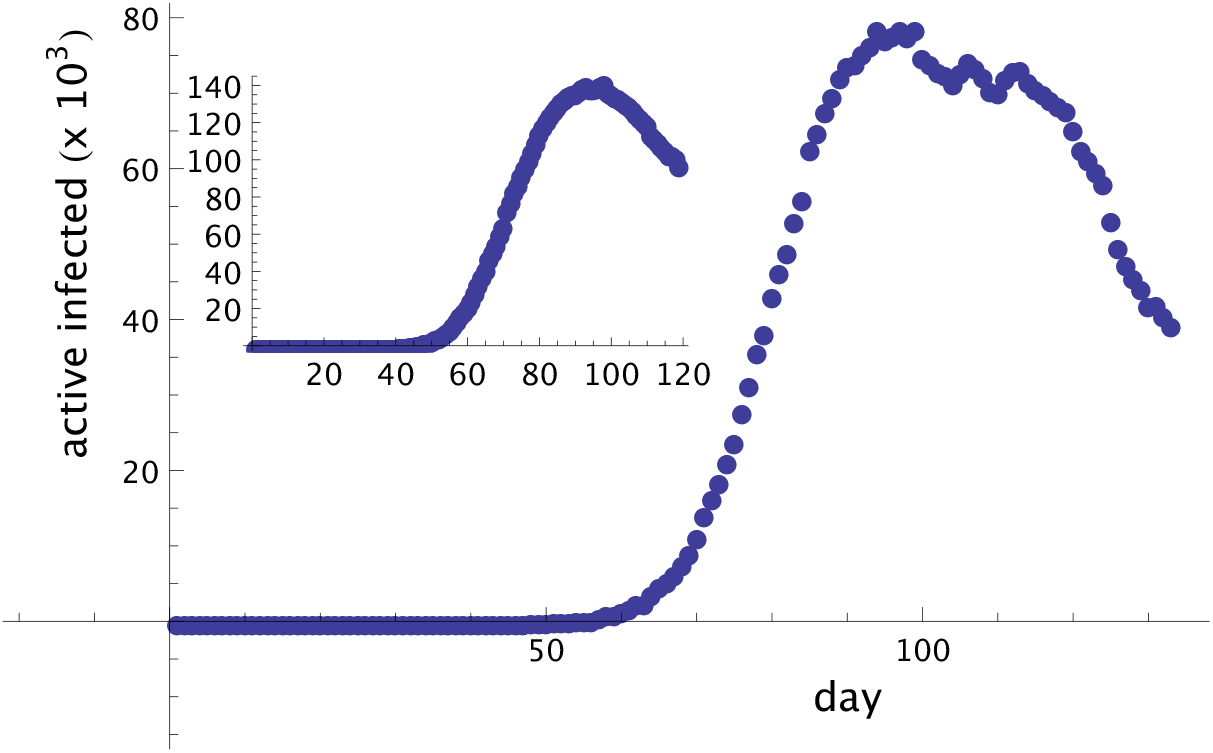
Number of symptomatic infected people, *I*_*UK*_, in UK, from January 17th until May 28th. In the inset the asymptomatic (but infected) people, *A*_*UK*_, is shown.

**Figure 4:**
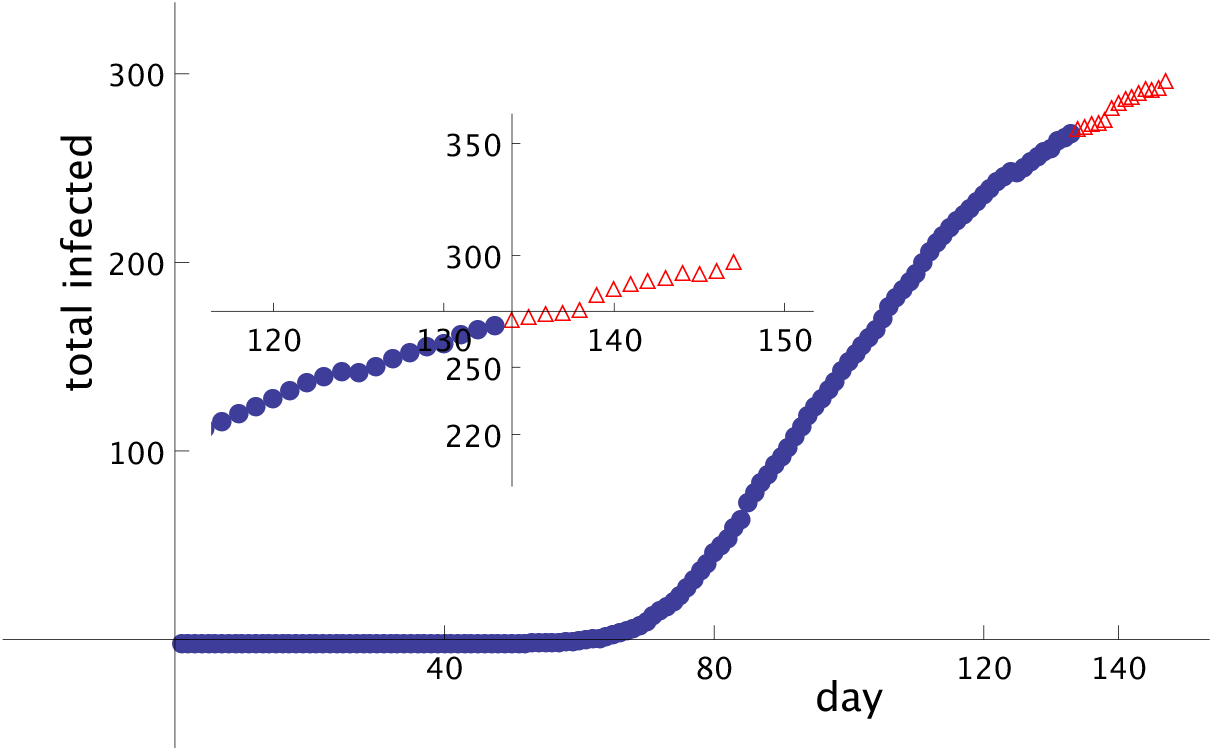
Total number of Covid-19 infected people in UK as function of the number of days, starting from January 17th, marked as number 1. Circle (blue) points are real data known up to day 133 (May 28th), last day where the data were collected, and empty triangles (red) points are the prediction of the model until day 147 (June 11th). *α* = 0.94 and *β* = 0.001. The inset shows a short scale, from day 117 up to day 147. The forecast (empty triangles, red) was made based on day 133 and previous days.

Therefore, to test the method in UK, we will consider our fictitious *t*_*N*_ = 129-th day, with four days less the true *t*_*N*_ = 133. As a consequence, it will be possible to test the days from 129 until 133, generated by the method (for this test we are considering only the real data of *I*_*UK*_(*t*) up to the day 129 and of *A*_*UK*_(*t*) up to day 115). The agreement among the real data and the points predicted by the method from day 129 until day 133 is quite good for *α* = 0.94 and *β* = 0.001 and the prediction points of the method follows the slope of the curve of total infected in UK. This procedure with these values for the parameters can then be applied to a real forecast from day 133 (the last day when we collected the data) until day 147 (June 11th). Fig. 4 show the real data with circles (blue) and the numbers predicted by the method with empty triangles (red) for *α* = 0.94 and *β* = 0.001. These numbers are shown in Table 2.

**Table 2:** Forecast of the total number of infected people by Covid-19 in UK from day 134 (May 29th) until day 147 (June 11th), see also Figure 4. Numbers are in thousands. This forecast was based on values of *I*_*UK*_ and *A*_*UK*_ until day 133 obtained from real data.

|  |  |  |  |  |  |  |  |
| --- | --- | --- | --- | --- | --- | --- | --- |
| day | 134 | 135 | 136 | 137 | 138 | 139 | 140 |
| $I_{tot}(t)$ | 273.0 | 274.4 | 275.7 | 276.5 | 277.8 | 284.2 | 286.9 |
| day | 141 | 142 | 143 | 144 | 145 | 146 | 147 |
| $I_{tot}(t)$ | 289.0 | 290.4 | 292.0 | 294.4 | 293.6 | 295.1 | 298.9 |

### 5.3 South Korea

The first infected person with symptoms in South Korea was identified on January 20th, our day *t*_0_. In our formalism this means that the first asymptomatic person was infected on January 6th, our day number one. The data set from January 06 (day 1) until May 28th (day 144), the last day were the data were collected, was obtained from the ECDC site [11], just to use another data base. Based on the data of total infected people from this site, we can construct the time series for the (symptomatic) infected people,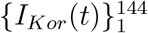, see equation (8). From the time series of {*I*_*Kor*_(*t*)}, and using equations (9) and (10), we can construct the time series for the asymptomatic people,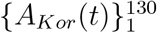. Figure 5 shows the graph of infected and asymptomatic people in South Korea in each day. In Figure 6 it is shown the real total number of infected people in South Korea until day 144, May 28th, with (blue) full circles. The empty triangles (in red) are the prediction of the method, keeping *α* = 1.04 and *β* = 0.02 constants, until day 158, June 11th. These forecast numbers are shown in Table 3

**Table 3:** Forecast of the total number of infected people by Covid-19 in South Korea from day 145 (May 29th) until day 158 (June 11th), see also Figure 6. Numbers are in thousands. This forecast was based on values of *I*_*Kor*_ and *A*_*Kor*_ until day 144 obtained from real data.

|  |  |  |  |  |  |  |  |
| --- | --- | --- | --- | --- | --- | --- | --- |
| day | 145 | 146 | 147 | 148 | 149 | 150 | 151 |
| $I_{tot}(t)$ | 11.381 | 11.412 | 11.454 | 11.490 | 11.506 | 11.539 | 11.565 |
| day | 152 | 153 | 154 | 155 | 156 | 157 | 158 |
| $I_{tot}(t)$ | 11.601 | 11.643 | 11.710 | 11.781 | 11.818 | 11.811 | 11.860 |

**Figure 5:**
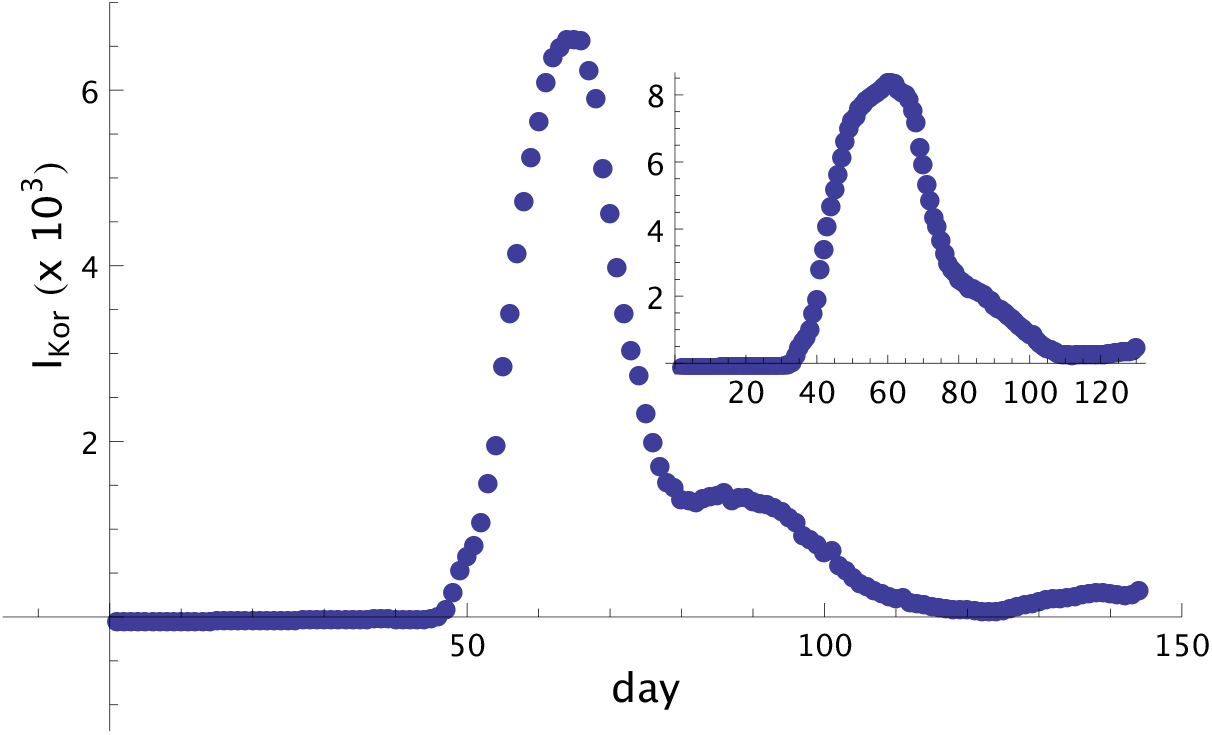
symptomatic infected (*I*_*Kor*_) people until day 144 (May 28th) in South Korea. Inset: asymptomatic (*A*_*Kor*_) people until day 126 (May 14th). The propagation of the disease is controlled but still some oscillation remain.

**Figure 6:**
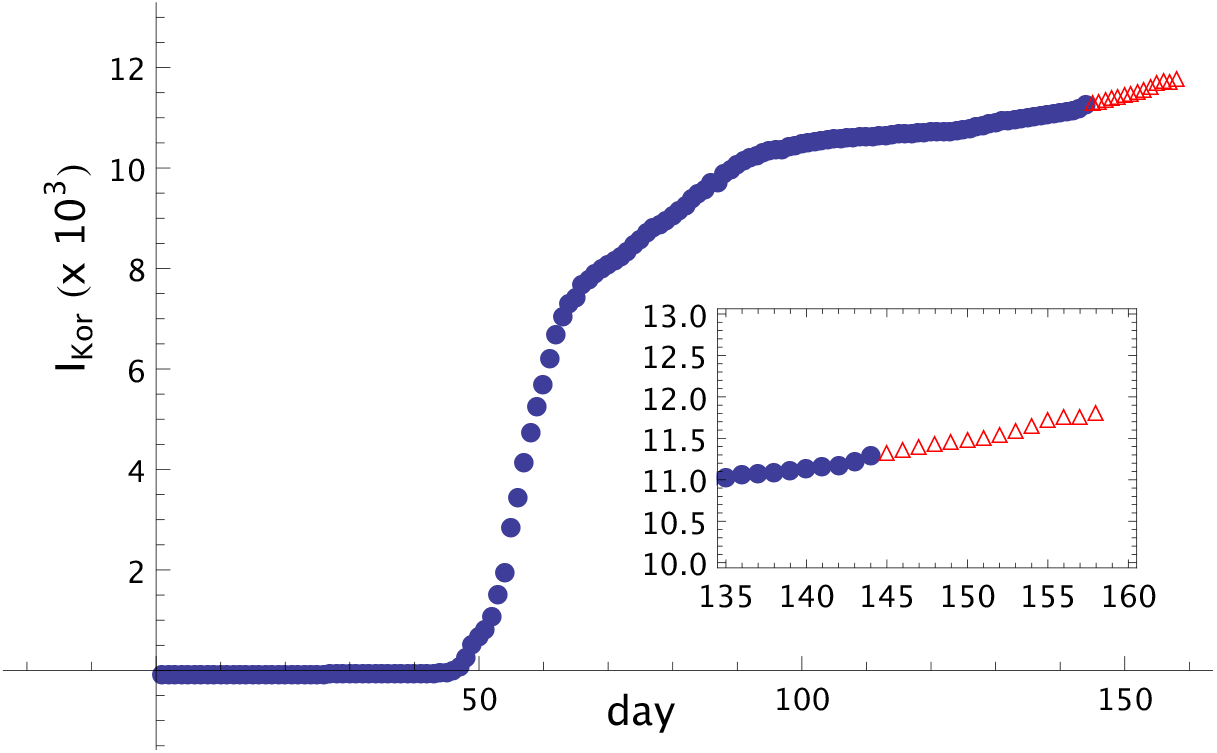
Total infected people in South Korea, (*I*_*Kor*_(*t*)), until day 144 (May 28th). Full circles (blue) are the real data from [11]. Triangles are the forecast, using the values *α* = 1.04 and *β* = 0.02, from day 144 until day 158 (June 11th). In the inset we can see the window from day 135 until day 158.

## 6 Discussion

In the present study we present a discrete-time model to study the time evolution of the Covid-19 pandemic based mainly on the numbers of asymptomatic and symptomatic infected people. The model has only two parameters, *α* and *β*. It is possible to use the daily updated data set of total confirmed cases of infected people to choose values for these parameters and predict how this number of infected people changes in the next 14 days if parameters are kept constant. The model gives the effective numbers of total infected people, in tens or hundreds of thousands, that can be directly compared with the data set available from distinct sources, including Johns Hopkins University, ECDC, Worldometer and others. Also, the model incorporates the average time-delay asymptomatic people take to become symptomatic people and the average time delay the symptomatic infected people take to recover or, eventually, die. Here, this model has been implemented in three countries (Brazil, UK and South Korea), and the fitting tested with past data indicates satisfactory accuracy, with error rates lower than 5% for the three tested countries. As such, we have found that the model is able to provide a reliable estimate of the status of the disease two weeks ahead of present time. Remarkably, this is achieved with a rather simple algorithm, based only on fitting two parameters and on data publicly available by distinct sources [5, 11, 12]. Altogether, we believe these characteristics facilitate its implementation by public sector, and may be a useful tool to track disease propagation and efficacy of public policies aiming to control it.

It may be argued that the model is over-simplistic and do not account for complexities from the pandemic evolution. For instance, it has been reported that the coronavirus can still be detected in throat swabs from recovered patients up to 13 days after they present no symptoms of the disease (as evaluated by clinical assessments), indicating that even people considered to be recovered from Covid-19 may still potentially infect others [13]. Furthermore, some patients may (eventually) quickly develop from asymptomatic phase *A* to recovered phase *R* without undergoing the symptomatic phase *I*. Finally, it has also been speculated the possibility of reinfection for patients who recovered from Covid-19 (in this regard, preliminary data from primates support the hypothesis of immunization and protection against reinfection [14]). In fact, we are aware that the model does not cover extensively the possible complexities related to disease propagation, but it focuses on the main course of propagation instead. As the model indicated a small error rate in its prediction, we considered it to fulfil its purpose adequately, exchanging complexity for simplicity in its comprehension and implementation while maintaining satisfactory predictive power (accuracy). These, in our understanding, are core principles for its assimilation by public health sector.

We would like to remark that the prediction for the total number of infected people is restricted to 14 days because of the definition given in equation (8), where we want to use the real data for the total number of infected people. If we relax this requirement to use real data, we can get points for the whole future, keeping the parameters constant. Certainly, this is not expected and this prediction for a long time is likely not reliable. It only gives an estimate of how will the disease propagate in the future if a given country does not change the value of the parameters. In fact, in practically all tested cases the parameters *α* and *β* decreased on time, likely reflecting measures against the propagation of the disease. For this reason, predictions provided by the model here presented are expected to decrease its accuracy for longer periods.

The people from phase *R* incorporates those who recovered as well as those who died following the infection. Thus, it is also possible to estimate separately the number of recovered and the number of dead people in two weeks, by simply applying in the *R* estimate the percentage of recovered and dead people from the studied country. As this is straightforward, we did not analyze this aspect in this study, but it is planned to be done in future works.

A last word about the model: it may also be used in other similar diseases - having both asymptomatic and symptomatic phases and both being contagious - as long as these other diseases have a daily (or some periodic) update of the data set. Clearly, the parameter *T* has to be adapted to the life cycle of the studied pathogen and it also is plausible to require two distinct *T* ^*I*^’s, one for the asymptomatic and other for the symptomatic phase. This, however, needs to be fitted to the specific characteristics of the studied disease.

Finally, a more extensive work is being prepared, with the analysis of many other countries and a study of the parameter space. We hope the present model can contribute to global efforts made to understand and control the Covid-19 pandemic.

## Data Availability

The data were obtained from Johns Hopkins University and ECDC

## Acknowledgements

EMFC acknowledges CNPq, FAPERJ and CAPES for financial support. MRC thanks INCT-SC for hospitality.

